# Heterologous boosting of neutralizing activity against Delta and Omicron SARS-CoV-2 variants in CoronaVac-primed adults; a randomized study with SCB-2019 vaccine

**DOI:** 10.1101/2022.12.06.22283103

**Authors:** Camilo C. Roa, Mari Rose A. de Los Reyes, Eric Plennevaux, Igor Smolenov, Branda Hu, Faith Gao, Hannalyn Ilagan, Donna Ambrosino, George Siber, Ralf Clemens

## Abstract

**Background:** The global COVID-19 pandemic has peaked but some countries such as China are reporting serious infectious outbreaks due to SARS-CoV-2 variants. Waning vaccine-derived immunogenicity and mutations in variants allowing vaccine evasion require new booster immunization approaches. We compared homologous and heterologous boosting in adults previously fully primed with a whole-virus inactivated COVID-19 vaccine.

**Methods:** At multiple sites in the Philippines we enrolled 430 adults (18-72 years) immunized with two doses of CoronaVac at least 3 months previously and randomly assigned them to receive homologous (CoronaVac, n = 216) or heterologous (recombinant protein vaccine, SCB-2019, n = 214) booster doses. Non-inferiority/superiority of the neutralizing antibody (NAb) response 15 days after boosting was measured by microneutralization against prototype SARS-CoV-2, and Delta and Omicron variants in subsets (50 per arm). Participants recorded solicited local and systemic adverse events for 7 days, unsolicited AEs until Day 29, and serious adverse events until Day 60.

**Results:** NAb geometric mean titers (GMT) against prototype on Day 15 were 744 (95% CI: 669-828) and 164 (143-189) in heterologous and homologous groups, respectively, with a heterologous/homologous GMT ratio of 4.63 (3.95-5.41), meeting both pre-defined non-inferiority and superiority criteria. Similarly, geometric mean-fold rises for NAb against Delta and Omicron BA.1, BA.2, BA.4 and BA.5 variants were superior after heterologous SCB-2019 (range 3.01-4.66) than homologous CoronaVac (range 0.85-1.6) in an exploratory analysis. Reactogenicity and safety measures were evenly balanced between groups; the most frequent local reaction was mild or moderate injection site pain; mild or moderate headache and fatigue were the most frequent systemic adverse events. No vaccine-related serious adverse events were reported.

**Conclusion:** Heterologous boosting of CoronaVac-immunized adults with SCB-2019 was well tolerated with superior immunogenicity than homologous boosting, particularly for newly emerged variants, supporting use of SCB-2019 for booster vaccination.

## BACKGROUND

Although the global peak appears to have passed, the COVID-19 pandemic continues to affect large numbers of people worldwide, notably with severe outbreaks ongoing in China due to the continuous emergence of new variants of the SARS-CoV-2 virus [1]. These new variants accumulate mutations in the primary antigenic target, the spike protein (S-protein), so that they evade immune protection induced by vaccines developed against the ancestral Wuhan-Hu-1 strain [2–4]. Decreases in vaccine-induced immunity due to the natural waning of antibody titers from the first month post-vaccination [5] and this lower susceptibility of the new variants to vaccine-induced immunity has led to the observation of lower protective efficacy from the first licensed COVID-19 vaccines [6,7]. This has driven the implementation of booster vaccination campaigns, especially with vaccines heterologous to the primary series in an effort to broaden the immune response and consequently the protection against new variants [8–13].

In the Philippines at the height of the pandemic a variety of COVID-19 vaccines were approved for use. One of these was the whole inactivated virus vaccine, CoronaVac, manufactured by SinoVac BioTech (Beijing, China). CoronaVac was approved by the WHO Strategic Advisory Group of Experts on Immunization (SAGE) for emergency use [14] and became one of the most widely used vaccines around the world. As part of a broader study of heterologous vaccination we investigated the impact of administering a booster dose of Clover Biopharmaceuticals’ COVID-19 vaccine candidate, SCB-2019, to CoronaVac-primed adults in the Philippines. SCB-2019 contains a pre-fusion form of the trimeric SARS-CoV-2 Spike protein (S-protein) derived from the ancestral strain (Wuhan-Hu-1) maintained in its trimeric structure using the proprietary Trimer-Tag® technology [15]. Here we report the interim results of that investigation including the immune responses against Delta and currently circulating Omicron SARS-CoV-2 variants to inform healthcare providers planning booster schedules in CoronaVac-primed populations.

## METHODS

This multi-center, observer-blind, randomized, controlled phase 3 study is ongoing in the Philippines to assess the immunogenicity and safety of SCB-2019 when administered as a heterologous booster dose to adults previously primed with a variety of licensed COVID-19 vaccines, in comparison with homologous booster doses of those vaccines. This interim report focuses on the comparison of the safety, reactogenicity and immunogenicity data following heterologous boosting with SCB-2019 with that following homologous boosting in adults primed with two doses of the whole inactivated virus vaccine, CoronaVac^®^. The study is registered on ClinicalTrials.gov (NCT05188677) and the protocol was approved by the Philippines Food and Drug Administration and all applicable institutional review boards. The study is being conducted in accordance with international guidelines including the Declaration of Helsinki and Council for International Organizations of Medical Sciences (CIOMS), and ICH GCP.

Eligible participants were male or female adults (≥ 18 years of age) with a documented history of two vaccinations with CoronaVac received at least three months previously. Main inclusion criteria were being healthy or having a stable pre-existing medical condition, being willing and able to comply with all study requirements and visits, and providing informed written consent to participate. Major exclusion criteria were any acute illness at the enrollment visit including a laboratory-confirmed SARS-CoV-2 infection initially detected by rapid antigen testing (RAT), or an axillary temperature ≥ 37.5°C, previous receipt of a COVID-19 vaccine other than CoronaVac (for this part of the study), any history of adverse events associated with vaccination or known allergy to any vaccine component, or receipt of any other investigational product within 30 days of study start.

### Vaccines

The investigational vaccine, SCB-2019, is manufactured by Clover Biopharmaceuticals (Changxing, China). Each 0.5 mL dose contains 30 μg SCB-2019 adjuvanted with 1.50 mg of the toll-like receptor agonist, CpG-1018 (Dynavax Technologies, Emeryville, CA, USA), and 0.75 mg aluminum hydroxide (Thousand Oaks Biopharmaceuticals, USA). Each dose of CoronaVac^®^ (SinoVac Life Sciences Co. Ltd., Beijing, China) contains 600 SU inactivated SARS-CoV-2 virus (CZ02 strain) and 0.225 mg aluminum hydroxide in 0.5 mL phosphate-buffered saline for injection. Both vaccines are administered by intramuscular injection in the deltoid of the non-dominant arm.

### Procedures

At enrollment on Day 1 participants were randomly allocated 1:1 using an interactive voice response system (IVRS), interactive web response system (IWRS), or interactive response technology (IRT) to two study groups and after an initial blood draw were administered either a homologous booster dose of CoronaVac or a heterologous booster dose of SCB-2019. Participants were blinded to which vaccine they received which was administered by study nurses who played no further role in the study. All subsequent steps, including laboratory analyses, were performed in a blinded manner. Participants were monitored for 30 minutes for any immediate reactions then recorded absence or occurrence with severity (mild, moderate, severe) of solicited local reactions and systemic adverse events (AE), and their axillary temperature daily for 7 days in an electronic study diary. Solicited local reactions were injection site pain, swelling and erythema; solicited systemic AEs were fatigue, headache, myalgia, arthralgia, loss of appetite, nausea, chills and fever (an axillary temperature ≥ 38°C). Unsolicited adverse events were recorded up to Day 29 and reported to the study investigator at each follow-up visit. Any serious adverse event (SAE), defined as death, or an AE that was life-threatening or led to hospitalization or persistent incapacity, or adverse events of special interest (AESI), defined as an AE potentially associated with COVID-19 or an autoimmune disorder, was to be reported immediately to the investigator throughout the study. Safety follow-up is ongoing for up to six months after vaccination. This interim report covers the first 60 days.

A second blood draw was performed on Day 15 to assess the immune response. Sera prepared immediately from both blood draws were stored at -80°C for shipping to VisMederi srl (Siena, Italy) for the immunogenicity analyses. Immunogenicity was measured in all available sera from Days 1 and 15 as virus neutralizing antibodies (NAb) in a microneutralization assay using prototype Wuhan-Hu-1 SARS-CoV-2 [10]. Neutralizing responses were also measured in a subset of each vaccine group (n = 50 per subset) against the SARS-CoV-2 variants, Delta (B.1.617.2) and Omicron sub-lineages BA.1, BA.2, BA.4, and BA.5.

### Statistics

The primary immunogenicity objective was to demonstrate non-inferiority of a heterologous SCB-2019 boost compared with a homologous CoronaVac boost, when measured as neutralizing response against prototype Wuhan-Hu-1 SARS-CoV-2. Non-inferiority would be met if the lower limit of the two-sided 95% confidence interval (CI) for the ratio of geometric mean titers (GMT) in the two groups (SCB-2019 GMT to CoronaVac GMT) was above 0.667 calculated using an ANCOVA model. Assuming a 5% drop-out, 212 enrolled subjects in each treatment group would provide 95.6% power to claim such non-inferiority of SCB-2019 booster to CoronaVac booster. When non-inferiority was confirmed, superiority was observed at the same time if the lower limit of the two-sided 95% CI for the ratio of GMT in the two groups (SCB-2019 GMT to CoronaVac GMT) was 1.5 or greater. The primary immunogenicity analysis was performed on all those who complied with the protocol and provided Day 15 blood samples within the prescribed time windows. Secondary immunogenicity objectives were to evaluate the neutralizing responses against SARS-CoV-2 variants in subsets of participants from homologous and heterologous groups. GMT ratios were assessed using an analysis of covariance (ANCOVA) model, including baseline antibody results and age as covariates, and vaccine treatment group as a fixed variable. Two-sided 95% CIs for GMT ratio were obtained by anti-log the confidence limits for the mean difference of the logarithmically transformed assay results which is calculated using t-distribution for the primary and the secondary immunogenicity objective analyses.

Responses are presented as GMTs at Days 1 and 15, geometric mean-fold rises (GMFR) from Day 1 to Day 15, and seroconversion rates (SCR) at Day 15 for all groups; seroconversion was defined as participants with a Day 1 titer below the lower limit of quantitation (LLoQ) having a Day 15 titer ≥ 4-fold LLoQ, or participants with a baseline titer above the LLoQ at Day 1 displaying a ≥ 4-fold increase in titer at Day 15. The SCR was the proportion (percentage) of each study group demonstrating seroconversion.

The reactogenicity and safety objectives in the current interim analysis were descriptive comparisons of incidence rates of solicited local reactions and systemic AEs through Day 7, unsolicited AEs up to Day 29, and SAEs and AESIs up to Day 60 in the two study groups. All participants who received their assigned dose of either vaccine (Safety Set) were included in the reactogenicity and safety analyses.

## RESULTS

This part of the study was conducted from June 13, 2022 to August 30, 2022. Following screening of 430 volunteers all were enrolled and randomly assigned to the CoronaVac (n = 216) and SCB-2019 (n = 214) groups. Overall, the mean age was 35.3 (± 11.6) years and there were proportionally more women than men (260 vs. 170). The mean (± SD) interval since their previous CoronaVac vaccination was 8.89 (± 2.11) months. These demographic characteristics were similar in the two study groups (**Table 1**). Following vaccination and checking of inclusion/exclusion criteria 420 participants were eligible for the Per Protocol immunogenicity analyses; 211 after homologous boosting with CoronaVac and 209 after heterologous boosting with SCB-2019 (**Figure 1**).

**Table 1.** Demographics of the exposed population (Safety Set)

| **Characteristics** | **Booster vaccine** |  |
| --- | --- | --- |
|  | **CoronaVac**<br>(N = 216) | **SCB-2019**<br>(N = 214) |
| **Sex – n, (%)** |  |  |
| Male | 76 (35.2) | 94 (43.9) |
| Female | 140 (64.8) | 120 (56.1) |
| **Mean age ± SD – years** | 35.5 ± 11.4 | 35.0 ± 11.9 |
| Median | 34.0 | 33.0 |
| Range | 18, 71 | 18, 72 |
| 18–59 years inclusive | 209 (96.8) | 206 (96.3) |
| ≥ 60 years | 7 (3.2) | 8 (3.7) |
| **Ethnicity – n, (%)** |  |  |
| Not Hispanic or Latino | 216 (100) | 214(100) |
| **Mean Body Mass Index ± SD – (kg/m<sup>2</sup>)** | 25.0 ± 5.3 | 24.4 ± 4.9 |
| **Time since last COVID-19 vaccination – months** |  |  |
| Mean ± SD | 8.95 ± 2.00 | 8.82 ± 2.22 |
| Range | 3.52, 13.8 | 3.45, 13.5 |

**Figure 1.**
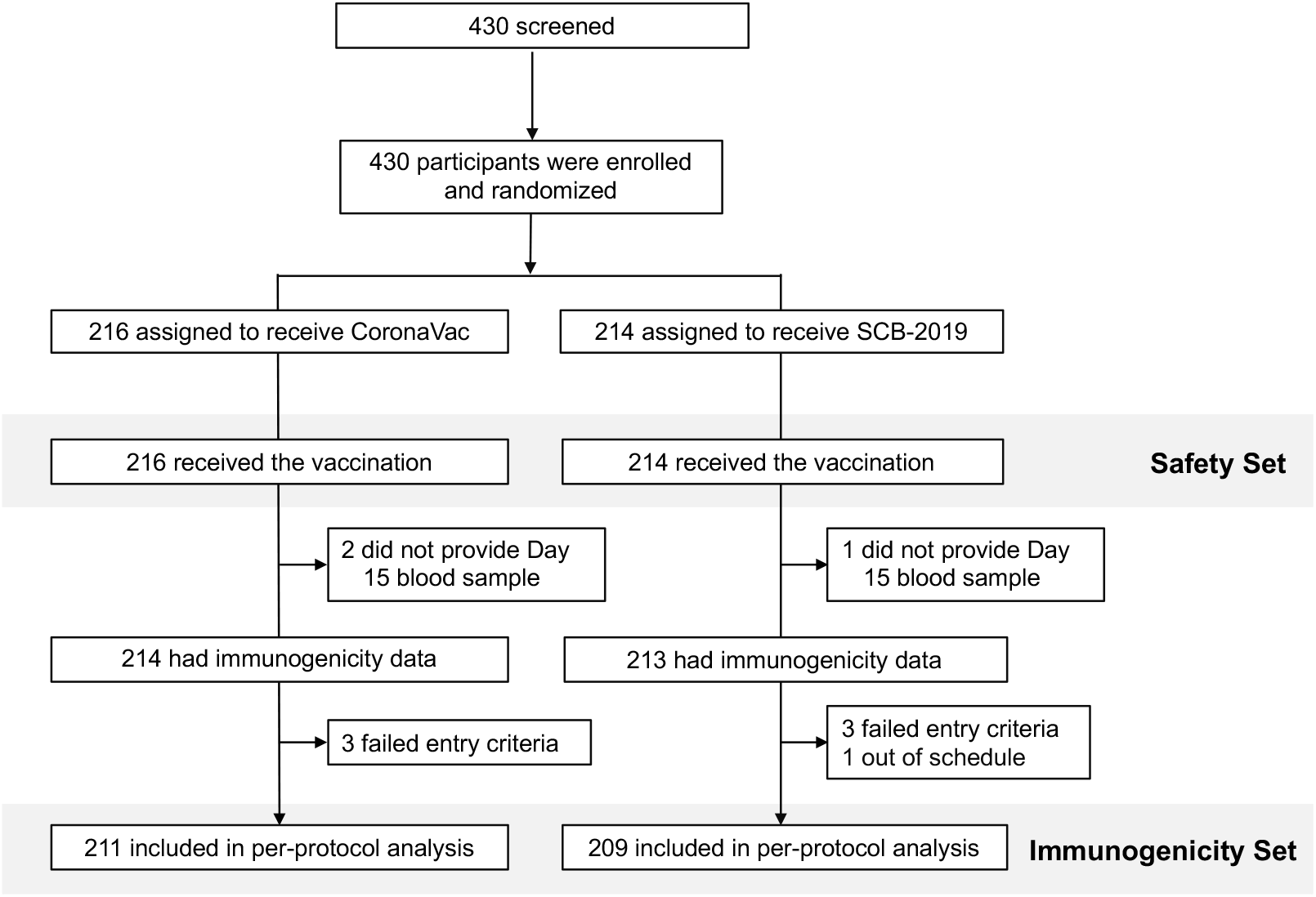
Disposition of participants in Safety Set and Per Protocol Immunogenicity Set

### Immunogenicity

Baseline immunogenicity was comparable in the two groups. Following boosting the primary immunogenicity objective was met. At Day 15 neutralizing antibody GMTs against prototype SARS-CoV-2 were 744 MN_50_ (95% CI: 669–828) and 164 MN_50_ (95% CI: 143–189) in heterologous (SCB-2019) and homologous (CoronaVac) groups, respectively, with a GMT ratio (GMT SCB-2019 / GMT CoronaVac) of 4.63 (95% CI: 3.96–5.41). As the lower 95% limit was greater than 0.667 non-inferiority was confirmed. Further, the lower 95% limit of the GMT ratio was greater than 1.5 showing this response was superior to that observed with a homologous CoronaVac booster (**Figure 2**).

**Figure 2.**
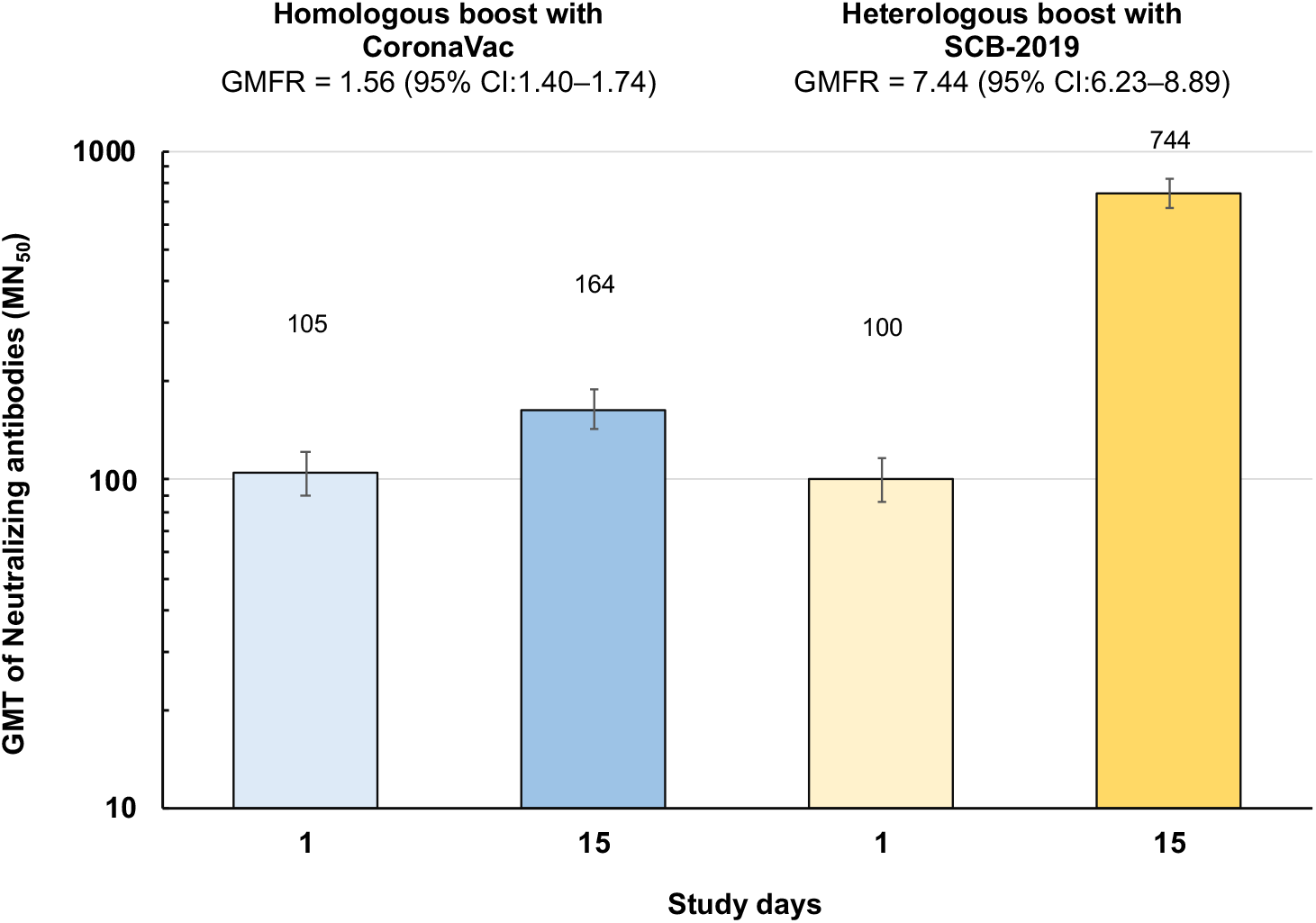
Immune responses in the two study groups as geometric mean titers (GMT) with 95% CI of neutralizing antibodies against prototype SARS-CoV-2 for 211 and 209 participants in CoronaVac and SCB-2019 groups on Day 1 and Day 15. Numbers in or above columns are GMT values.

Seroconversion rates at Day 15 also reflect the low neutralizing antibody response to the homologous booster with only 13.3% seroconverting against the prototype virus, compared with 71.3% of the heterologous group (**Table 2**). When assessed in the subsets of participants fewer than 5% seroconverted against Delta and the Omicron variants after homologous boosting compared with 60% against Delta and 34–62% against the various Omicron variants in the heterologous group. The GMTs and the GMFR achieved against these variants were consistently higher after heterologous boosting with SCB-2019 than those with the homologous CoronaVac booster (**Figure 3, Table 3**). In an exploratory analysis the GMT ratios for SCB-2019/CoronaVac were greater than 3.1 for all five variants tested with the lower 95% CI ranging from 2.45 to 2.97 (**Table 3**), demonstrating superiority of the heterologous response over the homologous response was observed for all five variants.

**Table 2.** Seroconversion rates for neutralizing antibodies (Per Protocol set and subsets)

| Assay |  | Booster vaccine |  |
| --- | --- | --- | --- |
|  |  | CoronaVac | SCB-2019 |
| Seroconversion for prototype SARS-CoV-2 – Per protocol set |  |  |  |
|  |  | (N = 211) | (N = 209) |
|  | n (%) | 28 (**13.3**) | 149 (**71.3**) |
|  | [95% CI] | [9.0–18.6] | [64.6–77.3] |
| Seroconversion for SARS-CoV-2 prototype and variants in subsets |  |  |  |
|  |  | (N = 49) | (N = 50) |
| Prototype | n (%) | 5 (**10.2**) | 33 (**66.0**) |
|  | [95% CI] | [3.4–22.2] | [51.2–78.8] |
| Delta | n (%) | 2 (**4.1**) | 30 (**60.0**) |
|  | [95% CI] | [0.5–14.0] | [45.2–73.6] |
| Omicron BA.1 | n (%) | 2 (**4.1**) | 28 (**56.0**) |
|  | [95% CI] | [0.5–14.0] | [41.3–70.0] |
| Omicron BA.2 | n (%) | 0 (**0**) | 28 (**56.0**) |
|  | [95% CI] | [0–7.3] | [41.3–70.0] |
| Omicron BA.4 | n (%) | 0 (**0**) | 17 (**34.0**) |
|  | [95% CI] | [0–7.3] | [21.2–48.8] |
| Omicron BA.5 | n (%) | 1 (**2.0**) | 31 (**62.0**) |
|  | [95% CI] | [0.1–10.9] | [47.2–75.3] |

**Table 3.** Neutralizing antibodies in the subsets to SARS-CoV-2 variants (MN_50_) and GMT ratio

|  |  | Booster vaccine |  | GMT ratio [95% CI]* |
| --- | --- | --- | --- | --- |
|  |  | CoronaVac | SCB-2019 | SCB-2019/CoronaVac |
| Variant |  | (N = 49) | (N = 50) |  |
| Prototype | **GMT** | **161.0** | **810.5** | **5.03** |
|  | [95% CI] | [129.4–400.5] | [652.5–1007] | [3.70–6.85] |
| Delta | **GMT** | **170.4** | **538.4** | **3.16** |
|  | [95% CI] | [145.3–199.9] | [460.0–630.4] | [2.52–3.96] |
| Omicron BA.1 | **GMT** | **67.1** | **223.0** | **3.33** |
|  | [95% CI] | [54.0–83.4] | [179.9–276.6] | [2.45–4.51] |
| Omicron BA.2 | **GMT** | **141.5** | **535.7** | **3.78** |
|  | [95% CI] | [119.1–168.5] | [451.6–635.5] | [2.97–4.83] |
| Omicron BA.4 | **GMT** | **42.5** | **154.9** | **3.64** |
|  | [95% CI] | [35.5–50.9] | [129.6–185.1] | [2.83–4.69] |
| Omicron BA.5 | **GMT** | **73.3** | **253.3** | **3.45** |
|  | [95% CI] | [61.6–87.3] | [213.2–301.0] | [2.70–4.42] |

\* GMT and GMT ratio calculated using an ANCOVA model.

**Figure 3.**
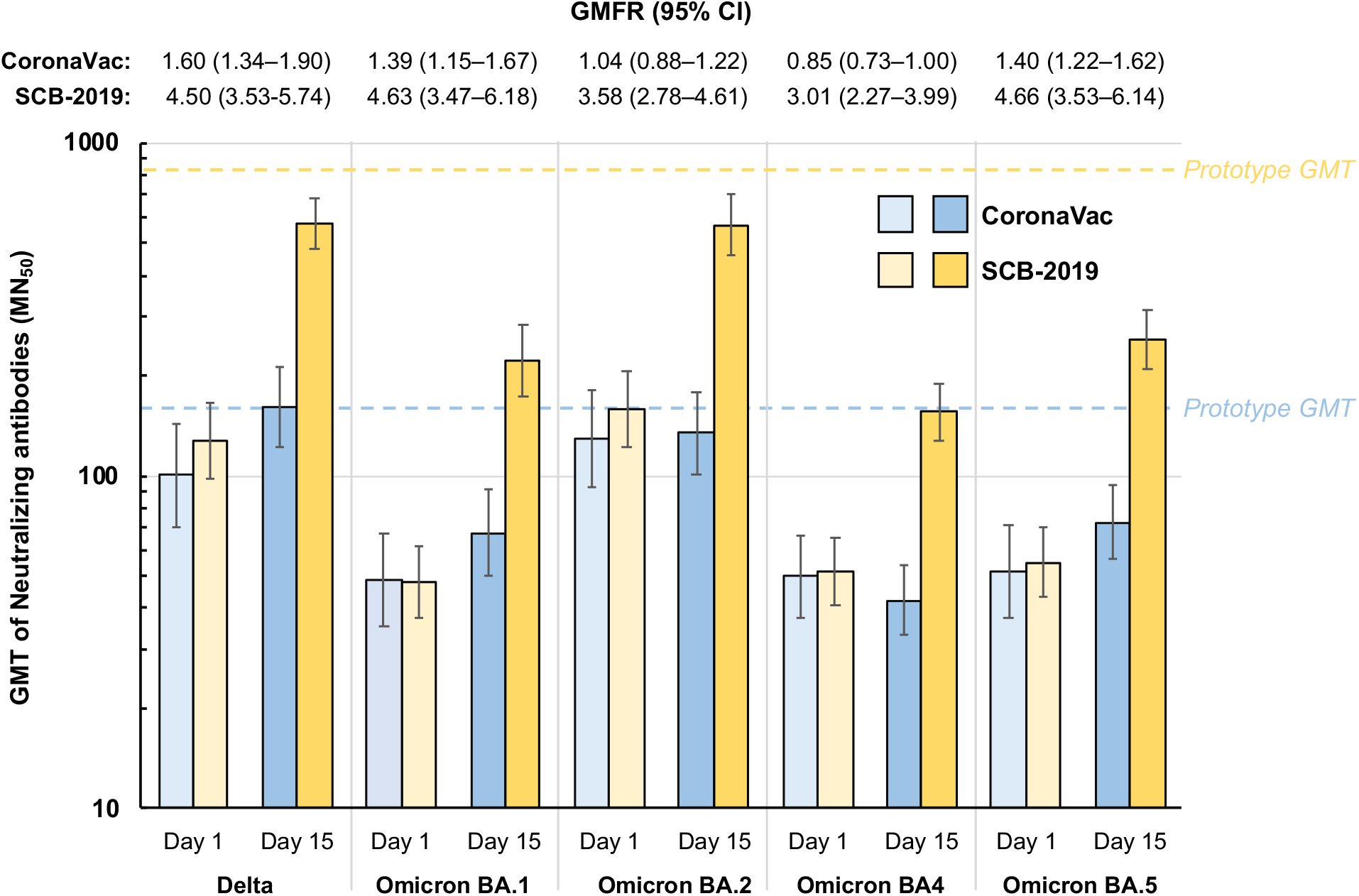
Immune responses as geometric mean titers (GMT) of neutralizing antibodies against Delta and Omicron SARS-CoV-2 variants in subsets of the CoronaVac (n = 49) and SCB-2019 (n = 50) study groups, with GMFR (95% CI) between Days 1 and 15 indicated. Dashed lines indicate the GMTs against the prototype SARS-CoV-2 virus in these SCB-2019 (upper line) and CoronaVac (lower line) booster subsets.

### Safety and reactogenicity

There were no immediate reactions to either vaccine and no deaths, nor any withdrawals or early study terminations due to an adverse event or AESI up to the Day 60 cut-off for this report. One SAE was reported, an acute kidney injury in the homologous group, which was not considered to be related to the vaccination by the investigator. Medically-attended adverse events were balanced between the two groups–6 events in 5 participants in the homologous group and 5 events in 4 participants in the heterologous group–and none were related to vaccination. Unsolicited adverse events were reported at similar rates in both groups, 36 events in 28 (13.0%) CoronaVac recipients and 33 events in 21 (9.8%) SCB-2019 recipients, three of which were considered to be related to vaccination in each group. Local reactions consisting exclusively mild or moderate pain at the injection site (**Figure 4**) were more frequent in the heterologous group (18.7%) than the homologous group (10.6%), but these were transient and resolved rapidly within the surveillance period. Systemic adverse events were reported by 21.5% and 18.5% of heterologous and homologous groups, respectively. The most frequent were fatigue and headache in both groups, the only severe cases being exclusively reported in the homologous group.

**Figure 4.**
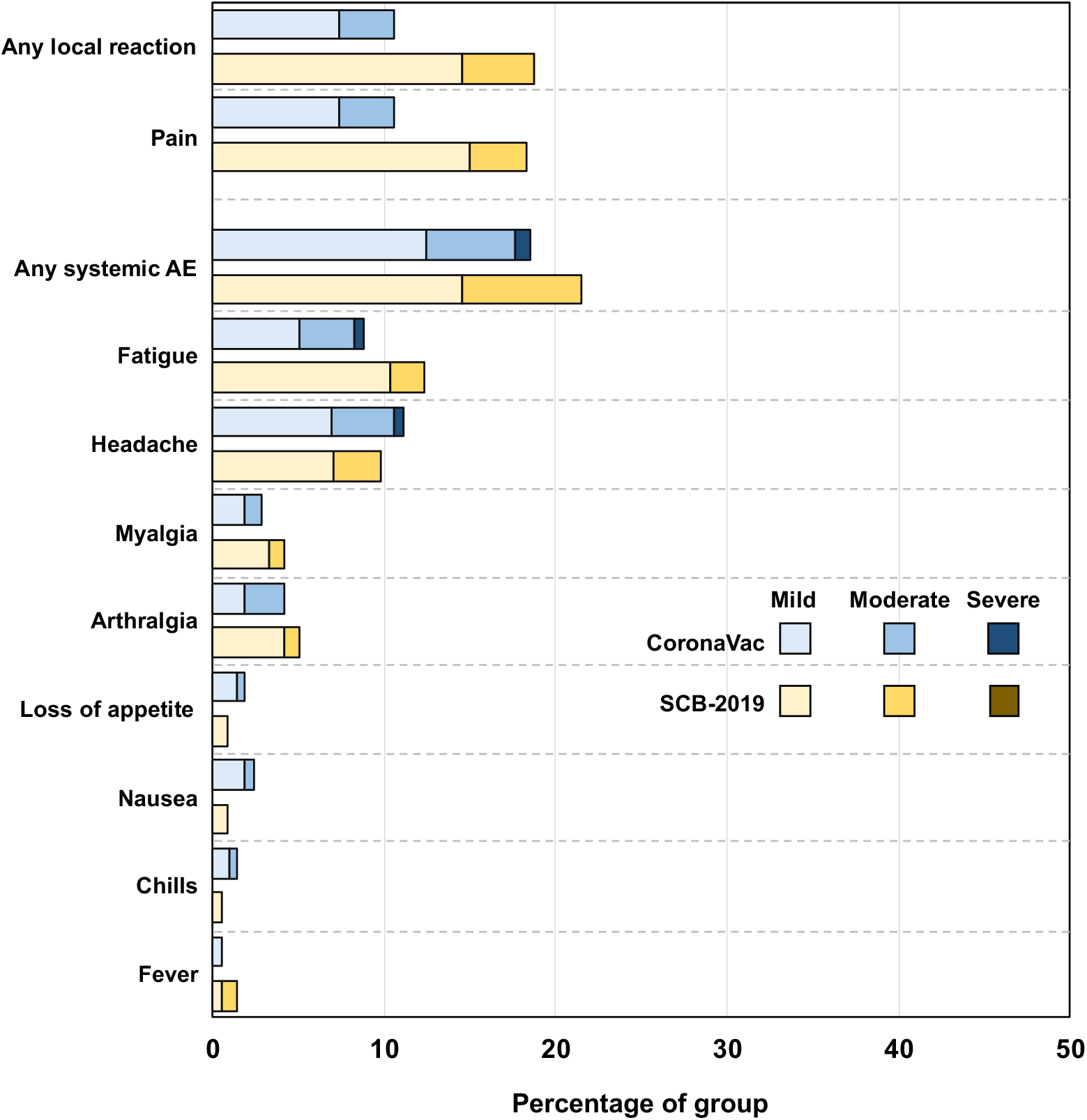
Proportions of the two study groups with solicited local reactions and systemic adverse events, with severity.

## DISCUSSION

This interim report of the heterologous boosting of immunity against SARS-CoV-2 with SCB-2019 vaccine in adults primed with two doses of CoronaVac at least three months earlier confirms not only are responses to heterologous boosting non-inferior to those after homologous boosting, but are actually superior. All the various immunogenicity parameters measured including GMFR, SCR and GMTs of neutralizing antibodies against SARS-CoV-2 variants show that heterologous boosting with SCB-2019 elicits a superior response than homologous boosting in CoronaVac-primed adults. This was achieved with no clinically meaningful differences in reactogenicity following either SCB-2019 or CoronaVac boosters, as all assessed safety and reactogenicity parameters generally occurred at low rates, were mainly mild or moderate in severity and were equally balanced between the two study groups.

In the current environment of continuing COVID-19 infections due to circulating new variants of SARS-CoV-2, predominantly the Omicron BA strains, the most important observation of this investigation of heterologous vs homologous boosting is the marked and consistent superior effect that heterologous boosting has on neutralizing activity against such variants. A homologous boost with CoronaVac did not elicit a consistent response against any of the variants tested, with a GMFR of 1.6 against Delta, and GMFRs ranging from 0.85 to 1.4 against Omicron BA.1, BA.2, and BA.5; the GMT against Omicron BA.4 was lower post-booster. In marked contrast, SCB-2019 elicited at least a three-fold increase in GMT against all five tested variants, with GMFRs ranging from 3.01 to 4.66.

A previously reported study investigated heterologous boosting with SCB-2019 in Brazilian adults primed at least 6 months previously with the ChAdOx1-S vector COVID-19 vaccine with immunogenicity assays performed in the same laboratory as the present study [10]. Heterologous SCB-2019 was more immunogenic than homologous ChAdOx1-S; 15 days after boosting with SCB-2019 or ChAdOx1-S the respective neutralizing antibody GMTs were 822 vs 274 against prototype SARS-CoV-2, 419 vs. 123 against Delta, and 65 vs. 31 against Omicron (B.1.1.529). The titers post-SCB-2019 boosting were consistent with those measured in the SPECTRA efficacy study in which SCB-2019 was shown to elicit a protective efficacy of 67·2% (95·72% CI: 54·3–76·8) against any COVID-19 disease and 100% (97·86% CI 25·3–100) against severe COVID-19 including hospitalizations [14].

These efficacy estimates were obtained against a background of circulating SARS-CoV-2 variants, mainly Delta, Gamma and Mu, but the high immunogenicity we show against the Omicron variants in this study support the notion that heterologous boosting with SCB-2019 would be efficacious against those new variants too. Heterologous boosting has been extensively investigated with some of the many possible combinations of other COVID-19 vaccines including priming with mRNA vaccines and heterologous boosting with vector-based or whole inactivated (CoronaVac) vaccines and have generally resulted in higher immunogenicity [9−13]. Importantly, the higher responses with heterologous boosters includes improvements in neutralizing activity against the most recent circulating variants, the Omicron sub-lineages. A review of studies on Omicron responses after primary and booster vaccination found booster doses generally resulted in higher neutralizing titers and response rates compared with post-primary responses [17]. A Chinese study found that in adults primed with two doses of CoronaVac heterologous boosting with one dose of either an mRNA vaccine, an adenovirus-vectored vaccine or a recombinant protein vaccine was more immunogenic in all cases than a homologous dose of CoronaVac, particularly against Omicron [11]; the highest response was to the recombinant protein vaccine, reflecting the results of our study. Studies in Hong Kong [18] and Thailand [19] found significantly higher neutralizing antibody responses to heterologous booster doses of BNT162b2 or ChAdOx-1S in CoronaVac-primed adults than homologous third doses of CoronaVac which included higher responses against Beta, Delta and Omicron variants, as well as against prototype SARS-CoV-2.

Heterologous boosting has been reported to be associated with higher rates of some systemic adverse events, notably fever, myalgia, malaise and fatigue [20], but we did not observe this in our study in which the only difference between homologous and heterologous groups was a small increase in the rate of transient, mild to moderate injection site pain in the heterologous group.

A major limitation of our study is that it only reports the immediate immune response following booster vaccination. As previously noted, immunity begins to wane within one month of completion of a primary series [5,6]. A small study of boosting of mRNA-vaccine primed adults with adenovirus-vector vaccine suggests that heterologous boosting provides an initially higher and then more durable immune response in terms of maintained levels of antibody titers [21]. As yet, we have no information of the durability of the response to an SCB-2019 booster, which will only come from a planned longer-term follow-up of the present study. The immune responses observed are also probably due to some degree of hybrid immunity as there was no selection of participants based on absence of evidence of previous SARS-CoV-2 exposure, either serologically or from medical history of COVID-19 infection or potentially from COVID-19 infection during the course of the study. However, this is a real-world observation and reflects the situation in which booster doses will be used in most countries around the world. One current exception is mainland China, where the vast majority of the 1.4 billion population remains naïve to SARS-CoV-2 infection and inactivated vaccines have been widely used. However, when we conducted a sensitivity analysis in our study to evaluate booster responses in a subset of those with low antibody levels at baseline we observed that after SCB-2019 booster the neutralization GMTs in this subset were consistent with the whole per-protocol set. This suggests that SCB-2019 could play an important role as a heterologous booster for inactivated vaccines regardless of prior infection history.

## CONCLUSION

This study is part of an ongoing study of the impact of using SCB-2019 as a heterologous booster of a variety of different COVID-19 vaccines used as primary doses. We are presenting this interim report showing the superiority of boosting immunity in CoronaVac-immunized adults by SCB-2019, particularly to elicit higher neutralizing antibody responses against currently circulating Omicron variants, because CoronaVac is one of the most widely used COVID-19 vaccines in primary immunization campaigns around the world. Our results are important to inform healthcare providers of the optimal alternative vaccines to use for booster campaigns in CoronaVac-immunized populations.

## Data Availability

Once the study is completed the datasets, including the redacted study protocol, redacted statistical analysis plan, and individual participants data supporting the results reported in this article, will be available three months from initial request, to researchers who provide a methodologically sound proposal, at the discretion of the company governing body. The data will be provided after its de-identification, in compliance with applicable privacy laws, data protection and requirements for consent and anonymisation.

## ACKNOWLEDGEMENTS

We are grateful to all the participants who volunteered to be in this study, and to the staff at all the study centers for their invaluable support. We thank the Philippines Department of Health for the generous provision of CoronaVac vaccine for this study. Editorial assistance in the preparation of this article was provided by Dr. Keith Veitch of keithveitch communications (Amsterdam, the Netherlands) with the financial support of Clover Biopharmaceuticals.

## AUTHOR CONTRIBUTIONS

Eric Plennevaux, Igor Smolenov, Branda Hu, Faith Gao, Hannalyn Ilagan contributed to the conception, design, and conduct of the study, analysis and interpretation of data and drafting the article. Camilo C. Roa, Jr., Mari Rose A. de Los Reyes contributed to the acquisition of data and revising the article. Donna Ambrosino, George Siber, and Ralf Clemens participated in the analysis and interpretation of data and revision of the article. All authors gave their final approval of the version to be submitted.

